# Evidence Behind the Automation of Clinical Trial Statistical Programming: A Scoping Review of Technology Adoption, Validation Frameworks, and AI/ML Integration (2020–2025)

**DOI:** 10.64898/2025.12.24.25342988

**Authors:** Jaime Yan, Jason Zhang, Tingting Tian

## Abstract

**Background and Objectives:** Pharmaceutical organizations are investing heavily in automating clinical trial statistical programming, yet the empirical basis for these decisions remains unclear. No prior review has systematically assessed whether claimed automation benefits are supported by rigorous evidence. This scoping review asks: (RQ1) What automation technologies have been adopted (2020–2025), and what evidence supports their effectiveness? (RQ2) What is the quality of evidence for validation approaches? (RQ3) What are the critical evidence gaps and research priorities?

**Methods:** Scoping review following PRISMA-ScR guidelines (22-item checklist provided). Six sources searched (2020–2025): PubMed, Google Scholar, arXiv, and conference proceedings. GRADE methodology assessed evidence quality. Of 987 publications screened, 262 met inclusion criteria.

**Results:** We found a critical asymmetry between technology readiness and evidence quality. Tools have reached production maturity—pharmaverse packages reduced TLF time 15–25% (GRADE: Low), RED-Cap2SDTM achieved an estimated 75–85% SDTM conversion savings (GRADE: Low), and domain-specific LLMs reached 88–93% F1 (GRADE: Moderate)—yet only 16% of publications reported quantitative outcomes. Most strikingly, just 2.3% of 527 validation papers contained effectiveness data, and **zero RCTs comparing validation approaches exist**. Double programming, the field’s most resource-intensive activity, rests on Very Low quality evidence.

**Conclusions:** Automation technology is mature, but the evidence base is predominantly Low to Very Low, creating risk that platform transitions are guided by anecdote rather than data. We propose a four-dimensional Automation Maturity Framework and a prioritized research agenda to address critical evidence gaps.

**Data Access Statement:** This study is a literature review synthesizing published research. All data supporting this review are derived from publicly available publications cited in the reference list.

## 1 Introduction

### 1.1 The Problem: Decisions Without Evidence

Clinical trial statistical programming, encompassing CDISC-compliant dataset creation (SDTM, ADaM), tables/listings/figures (TLF) generation, and validation, is undergoing a rapid transformation from manual SAS^®^-based workflows toward metadata-driven, automated pipelines. A typical Phase III trial requires 200–500 TLFs, each validated through independent double programming that consumes 1.6–2.0× the primary programming effort and extends development timelines by 20–40%^1^. As organizations invest millions in multi-year platform transitions (e.g., SAS to R/Python), they face an important question: *are these investment decisions supported by empirical evidence, or merely by vendor claims and anecdotal reports?*

This question matters because the stakes are substantial. Median Phase III trial costs reach $19M^1^, data complexity is rising from decentralized trials and real-world evidence integration, and regulatory agencies increasingly accept (and sometimes expect) automated, reproducible workflows. Yet when we examined the literature, we found no prior systematic evidence synthesis evaluating whether claimed efficiency gains in *statistical programming* automation are supported by quantitative effectiveness data. Previous reviews addressed general data management transformation^2^ and administrative AI applications in clinical trials^3^, but none specifically assessed the *evidence quality* behind statistical programming automation claims. This gap means organizations may be making consequential technology adoption decisions based on anecdote rather than data.

### 1.2 Rationale: Why This Review Is Needed Now

Three converging developments in 2022–2024 have created urgency for evidence-based guidance:

1. **Regulatory paradigm shift**: The FDA’s Computer Software Assurance (CSA) guidance (September 2022) formally endorsed risk-based approaches over exhaustive testing; the ICH E6(R3) Step 2 draft (May 2023) explicitly addressed electronic systems and risk-proportionate quality management; and the R Consortium completed five successful FDA pilot submissions (2021–2024), establishing precedent for open-source regulatory packages^6,16^.
2. **Standards maturation**: The CDISC Analysis Results Standard (ARS) v1.0 (April 2024) provided the first machine-readable specification for TLF generation, enabling a fundamentally new automation paradigm^7^.
3. **AI disruption**: Large language models (2023–2025) introduced code generation capabilities that challenge existing validation assumptions, yet no regulatory framework addresses AI-assisted programming in clinical contexts^20,23^.

These developments create both opportunity and risk. Organizations that adopt automation prematurely, without understanding the evidence base, may compromise regulatory compliance. Organizations that delay adoption unnecessarily may lose competitive advantage. What is needed is not another catalog of available tools, but a rigorous assessment of the *evidence* supporting those tools. This review addresses that need by asking not only what has been developed, but whether it *works as claimed*, and where the evidence falls short.

### 1.3 Objectives and Research Questions

This scoping review addresses three research questions using the Population–Concept–Context (PCC) framework, each designed to move beyond cataloging tools toward assessing the evidence that supports them:

- **RQ1 (Technology and evidence):** What automation technologies have been developed or adopted for clinical trial statistical programming between 2020–2025 (Population: statistical programmers and clinical data managers; Concept: automation tools for TLF generation, CDISC dataset creation, and validation; Context: pharmaceutical and biotech industry, 2020–2025), and what empirical evidence supports their *claimed* effectiveness?
- **RQ2 (Validation evidence quality):** What is the quality and quantity of empirical evidence— assessed using the GRADE framework—supporting current validation approaches (double programming, risk-based, automated testing)? This question was motivated by the observation that validation is the single most resource-intensive activity in statistical programming, yet its effectiveness has never been systematically evaluated.
- **RQ3 (Research priorities):** What are the critical evidence gaps, and what specific study designs, sample sizes, and outcome metrics are needed to generate the actionable evidence the field currently lacks?

Rather than simply inventorying available tools, we use these questions to reveal a central finding: the asymmetry between technology maturity and evidence quality. To synthesize this insight, we propose a four-dimensional **Automation Maturity Framework** (AMF)—spanning technology readiness, evidence quality, regulatory acceptance, and organizational adoption—that enables practitioners to assess not just whether a tool *exists*, but whether its adoption is *supported by evidence*.

## 2 Methods

### 2.1 Study Design and Protocol Registration [PRISMA-ScR Items 1, 5]

This scoping review follows the PRISMA-ScR (Preferred Reporting Items for Systematic Reviews and Meta-Analyses extension for Scoping Reviews) guidelines^47^; the completed 22-item PRISMA-ScR checklist with page and line references is provided in Supplementary Table S1. The title identifies this as a scoping review (PRISMA-ScR Item 1). The protocol was registered on the Open Science Framework (OSF; registration DOI: [to be inserted upon registration]). We acknowledge retrospective registration as a limitation and discuss its implications in Section 4.5. The study selection process is illustrated in the PRISMA flow diagram (Figure 1).

**Figure 1:**
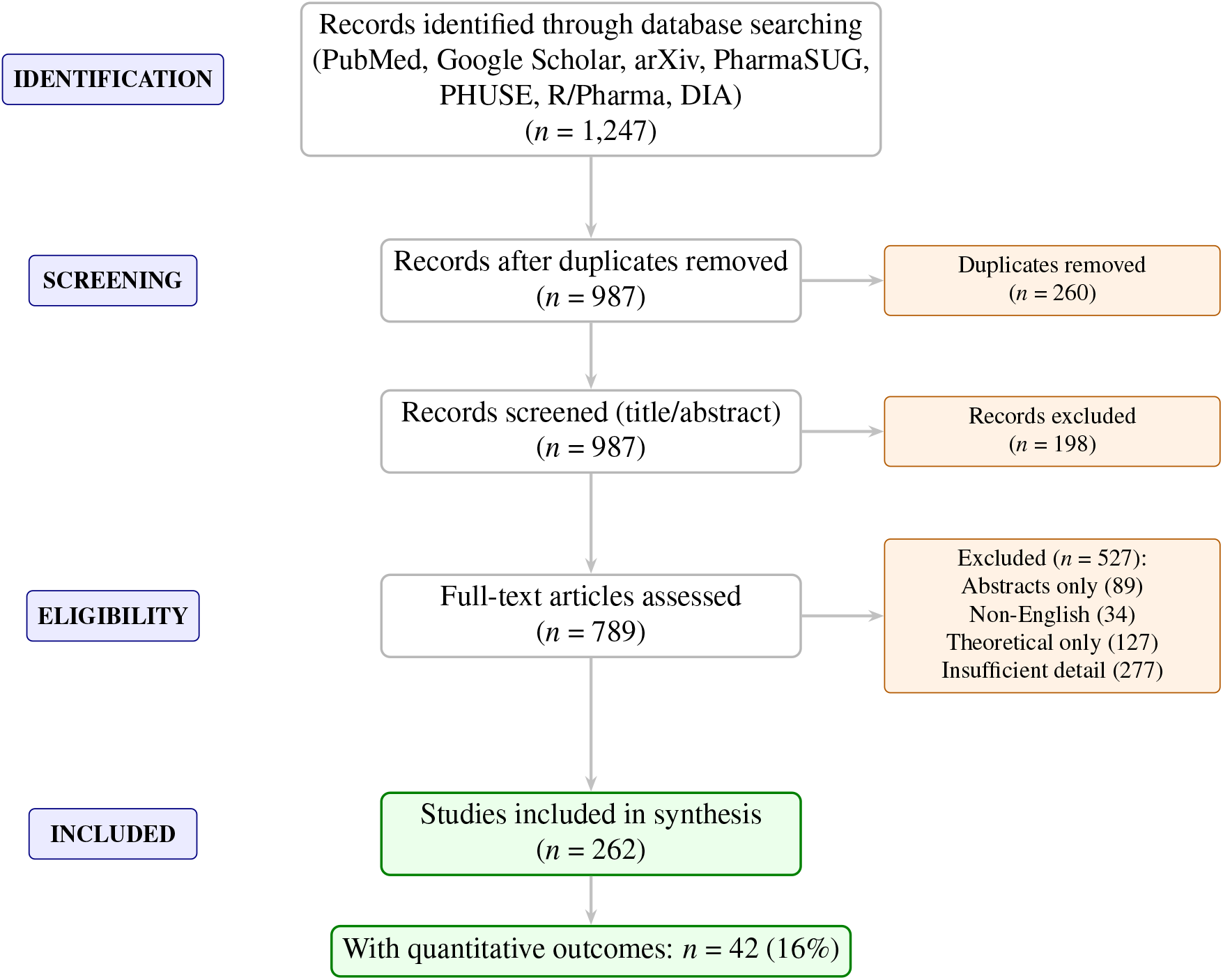
PRISMA flow diagram for study selection. From 1,247 initial records, 262 publications met inclusion criteria for qualitative synthesis, with 42 (16%) providing quantitative data suitable for synthesis.

### 2.2 Eligibility Criteria [PRISMA-ScR Item 6]

Eligibility was defined using the Population–Concept–Context (PCC) framework, consistent with JBI methodology for scoping reviews:

- **Population**: Statistical programmers, clinical data managers, and biostatisticians working in clinical trial programming
- **Concept**: Automation tools, frameworks, and methodologies for TLF generation, CDISC dataset creation (SDTM/ADaM), validation, and AI/ML-assisted code generation
- **Context**: Pharmaceutical, biotechnology, and contract research organizations conducting regulatory submissions (FDA, EMA, PMDA), 2020–2025

**Inclusion criteria** (1) Publications describing automation frameworks, validation strategies, or AI/ML applications relevant to clinical trial statistical programming; (2) published in English between January 2020 and December 2025; (3) containing sufficient technical detail for synthesis (implementation architecture, quantitative outcomes, or validated methodology). **Exclusion criteria:** (1) abstracts only without full text (excluded because insufficient detail for evidence quality assessment); (2) duplicate publications; (3) non-English language; (4) purely theoretical discussions without implementation details or empirical evidence; (5) publications focused exclusively on clinical data management without statistical programming content. Exclusion criteria were applied to ensure that included sources could contribute meaningfully to evidence quality assessment.

### 2.3 Information Sources and Search Strategy [PRISMA-ScR Items 7, 8]

We searched six information sources: PubMed (last searched December 15, 2025), Google Scholar, arXiv, and industry conference databases (PharmaSUG proceedings 2020–2025, PHUSE proceedings 2020–2025, R/Pharma proceedings 2020–2025, DIA proceedings 2020–2025). Additionally, we hand-searched reference lists of included studies and consulted CDISC, FDA, and EMA websites for regulatory guidance documents.

The search was conducted in two phases. Phase 1 (core review, January–March 2025) identified publications on automation frameworks. Phase 2 (validation meta-analysis, April–June 2025) screened additional validation-focused papers. The full electronic search strategy for PubMed is provided in Supplementary Table S2; briefly, the strategy combined MeSH terms and free-text keywords: (“clinical trial” OR “clinical study”) AND (“statistical programming” OR “SAS programming” OR “CDISC” OR “SDTM” OR “ADaM” OR “TLF” OR “tables listings figures”) AND (“automation” OR “metadata-driven” OR “validation” OR “double programming” OR “R programming” OR “pharmaverse” OR “artificial intelligence” OR “machine learning” OR “large language model”). Search strategies for all databases are available from the corresponding author upon request.

### 2.4 Selection of Sources of Evidence [PRISMA-ScR Item 9]

Two reviewers (JY, JZ) independently screened titles and abstracts against the eligibility criteria. Full-text screening was performed by JY with verification by TT for a random 20% sample. Disagreements were resolved through discussion; in cases where consensus could not be reached, all three authors reviewed the source collaboratively. Inter-rater agreement was not formally calculated, which we acknowledge as a limitation. The screening process yielded the following numbers: 1,247 records identified, 260 duplicates removed, 987 screened at title/abstract level, 198 excluded at screening, 789 assessed at fulltext level, and 527 excluded at full-text (reasons: abstracts only [*n*=89], non-English [*n*=34], theoretical only [*n*=127], insufficient detail [*n*=277]). These data are summarized in the PRISMA flow diagram (Figure 1).

### 2.5 Data Charting Process [PRISMA-ScR Item 10]

A standardized data charting form was developed in Microsoft Excel, pilot-tested on 15 publications, and iteratively refined. Data were extracted by JY and verified by JZ for quantitative entries. The charting process followed a three-step procedure: (1) independent extraction by JY using the standardized form; (2) verification of quantitative entries by JZ; (3) consensus resolution for any discrepancies through discussion.

### 2.6 Data Items [PRISMA-ScR Item 11]

The charting form captured the following data items for each included source:

- *Bibliographic*: Author(s), year, title, publication type (journal, conference, guidance, documentation), country/region
- *Methodological*: Study design (RCT, controlled evaluation, case study, before/after, expert opinion), sample size, comparator (if any)
- *Domain*: Primary topic (TLF generation, CDISC automation, validation, AI/ML, platform/architecture)
- *Technology*: Tools/platforms described, programming language(s), CDISC standard version(s)
- *Outcomes*: Quantitative metrics (time reduction %, error detection rate, F1/accuracy, cost savings) with variance estimates where reported; qualitative findings
- *Evidence quality*: GRADE rating with justification

**Sources:** Included publications comprised peer-reviewed journals (31%, *n*=81), conference proceedings (47%, *n*=123; PharmaSUG, PHUSE, R/Pharma, DIA), regulatory guidance documents (8%, *n*=21; FDA, EMA, ICH), and authoritative open-source package documentation and CDISC standards (14%, *n*=37).

### 2.7 Critical Appraisal of Evidence [PRISMA-ScR Item 12]

Although critical appraisal is optional in scoping reviews per PRISMA-ScR, we deliberately chose to apply the GRADE (Grading of Recommendations, Assessment, Development, and Evaluation) framework— adapted for technical literature—to address RQ2 (evidence quality assessment). This decision was motivated by the observation during preliminary screening that most efficiency claims lacked rigorous empirical support, making an evidence quality assessment not merely informative but essential. Evidence was rated as: *high* (RCTs or well-designed comparative studies with consistent results); *moderate* (controlled evaluations on benchmark datasets with standard metrics); *low* (observational studies, before/after comparisons without controls); or *very low* (case reports, expert opinion, single-organization implementations). Publication bias was qualitatively assessed through examination of outcome directionality across studies. The GRADE approach, while designed for clinical interventions, has been adapted for diverse evidence types including public health and health systems research^48^, supporting its extension to technical literature. Our adaptation substituted “controlled evaluation on benchmarks” for “well-designed cohort studies” in the Moderate category, reflecting that technical evaluations typically use standardized datasets and reproducible metrics rather than patient cohorts.

### 2.8 Synthesis of Results [PRISMA-ScR Item 13]

Of the 789 publications screened, 262 met the inclusion criteria for the core review. Topic distribution: TLF generation (28%, *n*=73), CDISC dataset automation (24%, *n*=63), validation frameworks (22%, *n*=58), AI/ML applications (15%, *n*=39), and platform/architecture (11%, *n*=29). Only 42 publications (16%) reported quantitative outcomes suitable for synthesis, while the remainder provided qualitative descriptions or implementation guidance. Results are organized by research question: RQ1 (technology and evidence) is addressed across Sections 3.1, 3.3–3.5; RQ2 (validation evidence quality) in Section 3.2; RQ3 (research priorities) is addressed in Section 4.1.3.

**Statistical methods:** Quantitative synthesis methods were planned *a priori* but limited by data availability:

- *Planned*: Random-effects meta-analysis (DerSimonian-Laird) for error detection rates; standardized mean differences (SMD) for efficiency metrics; I^2^ heterogeneity assessment; funnel plot and Egger’s test for publication bias; subgroup analyses by platform (SAS vs. R vs. Python) and endpoint risk level
- *Actual*: Narrative synthesis with descriptive statistics (ranges, medians) due to: (a) only 12 of 527 validation studies (2.3%) with any quantitative data, of which just four provided directly comparable approach-specific effectiveness data; (b) missing variance estimates (SD, SE, CI) in primary sources; (c) heterogeneous outcome definitions precluding pooling; (d) no RCTs identified
- *Power analysis*: Post-hoc calculation indicated that detecting a 10 percentage-point difference in error detection rates (80% power, *α*=0.05) would require approximately 200 programs per arm— far exceeding available data (*n*=15 in largest study)

Effect sizes were characterized qualitatively as: *large* (*>*20% difference), *moderate* (10–20%), or *small* (*<*10%). For AI/ML benchmarks with multiple studies, effect sizes were estimated using Cohen’s *d* where mean and SD could be extracted or approximated from reported ranges (assuming range/4≈SD for normally distributed data).

## 3 Results [PRISMA-ScR Items 14–17]

This section presents findings organized by research question. Section 3.1 (RQ1: Technology and Evidence) maps the state of automation across four domains—TLF generation, CDISC dataset automation, validation frameworks, and AI/ML integration—while critically assessing the evidence supporting each domain. Section 3.2 (RQ2: Evidence Quality) provides a focused analysis of the validation evidence gap. Characteristics of the 262 included studies are summarized in Table 6 (PRISMA-ScR Item 15). The PRISMA flow diagram (Figure 1) documents the selection process (PRISMA-ScR Item 14).

### 3.1 RQ1: Technology Landscape and Evidence Quality

#### 3.1.1 TLF Generation Automation

Tables, Listings, and Figures (TLFs) constitute the primary evidence artifacts in clinical study reports. Key challenges include: specification complexity (intricate formatting, conditional logic, therapeutic area conventions); consistency requirements across studies; reproducibility for regulatory review; and quality control burden from traditional double programming^4^.

The TLFQC platform exemplifies emerging “codeless” approaches where users specify TLF requirements through graphical interfaces rather than programming, lowering barriers to automation adoption^36^.

Case studies report 15–25% reductions in TLF development time when transitioning from traditional SAS macro libraries to pharmaverse workflows^5^. Key advantages include: transparent open-source code enabling auditability; modular architecture supporting reuse; native integration with Git version control and CI/CD pipelines; and growing regulatory acceptance demonstrated through FDA and EMA submissions^6^.

**Critical assessment (RQ1):** While these reported gains are directionally encouraging, the evidence quality is Low (GRADE). The 15–25% figure derives from industry reports and conference presentations rather than controlled studies—no published study has compared pharmaverse workflows against SAS macro libraries in a randomized or matched design, and no variance estimates or confidence intervals have been reported. This means organizations cannot determine whether 15–25% is a reliable estimate or an optimistic outlier. Adoption estimates (e.g., SAS at 95%, R at 60%) derive from literature synthesis rather than systematic surveys, and likely reflect large-pharma bias in publication patterns.

#### 3.1.2 Metadata-Driven TLF Generation

A paradigm shift has occurred from code-centric to specification-centric TLF development (Figure 2). In metadata-driven approaches, TLF specifications are captured in structured formats (Excel, YAML, JSON), and generation engines interpret these specifications to produce outputs automatically.

**Figure 2:**
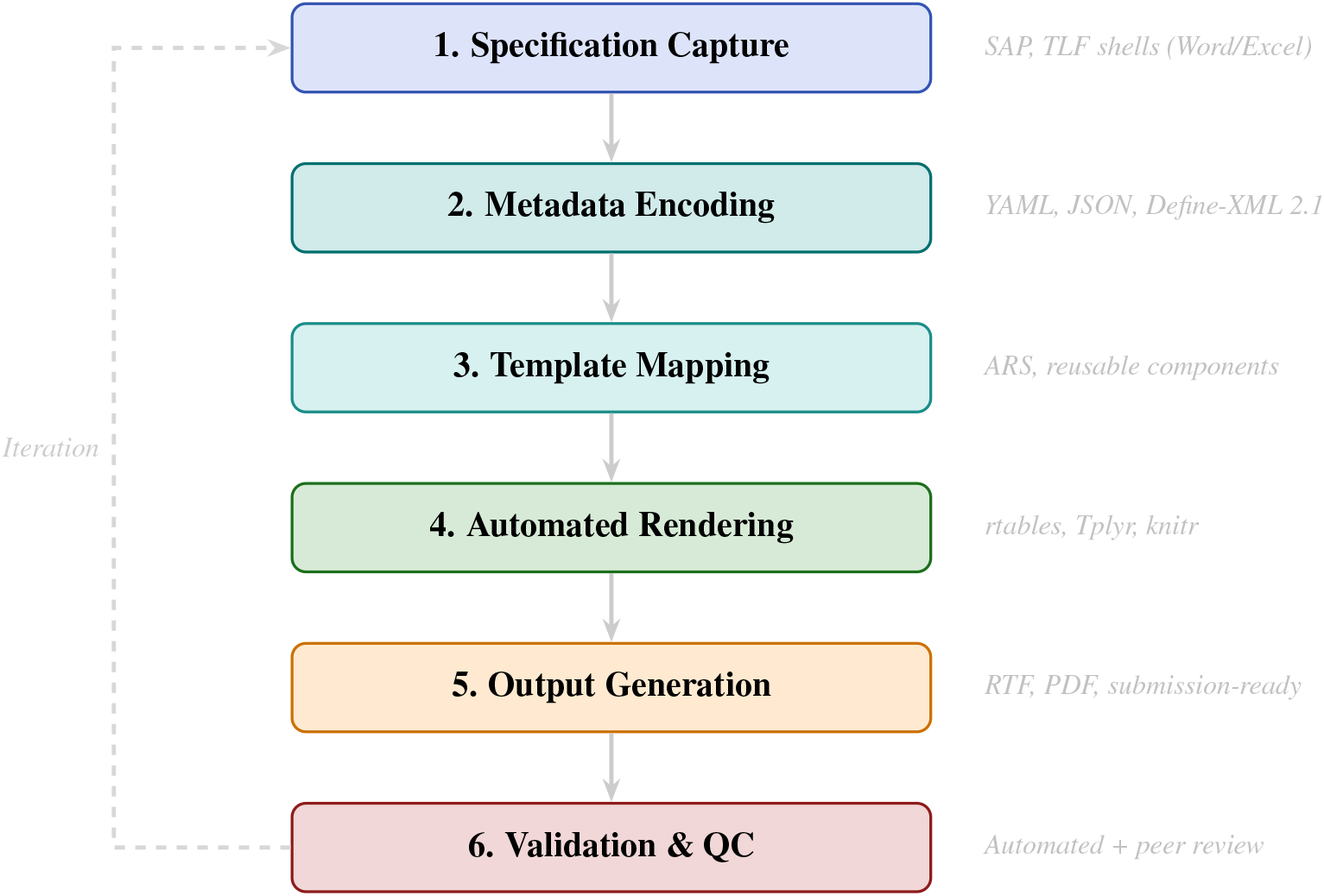
Metadata-driven TLF generation workflow. The six-step pipeline transforms SAP specifications into validated outputs, with feedback loops enabling iterative refinement. This approach reduces manual coding by 30–50% compared to traditional methods.

The CDISC Analysis Results Standard (ARS) provides a framework for machine-readable TLF specifications, enabling: automated TLF generation from rendering engines; explicit traceability between SAP, code, datasets, and outputs; reusability across studies; and regulatory transparency^7^. However, as of 2025, ARS adoption remains in early stages with limited production implementations.

#### 3.1.3 Quantitative Impact

Table 2 summarizes reported efficiency gains from TLF automation implementations across the literature.

**Table 1:**
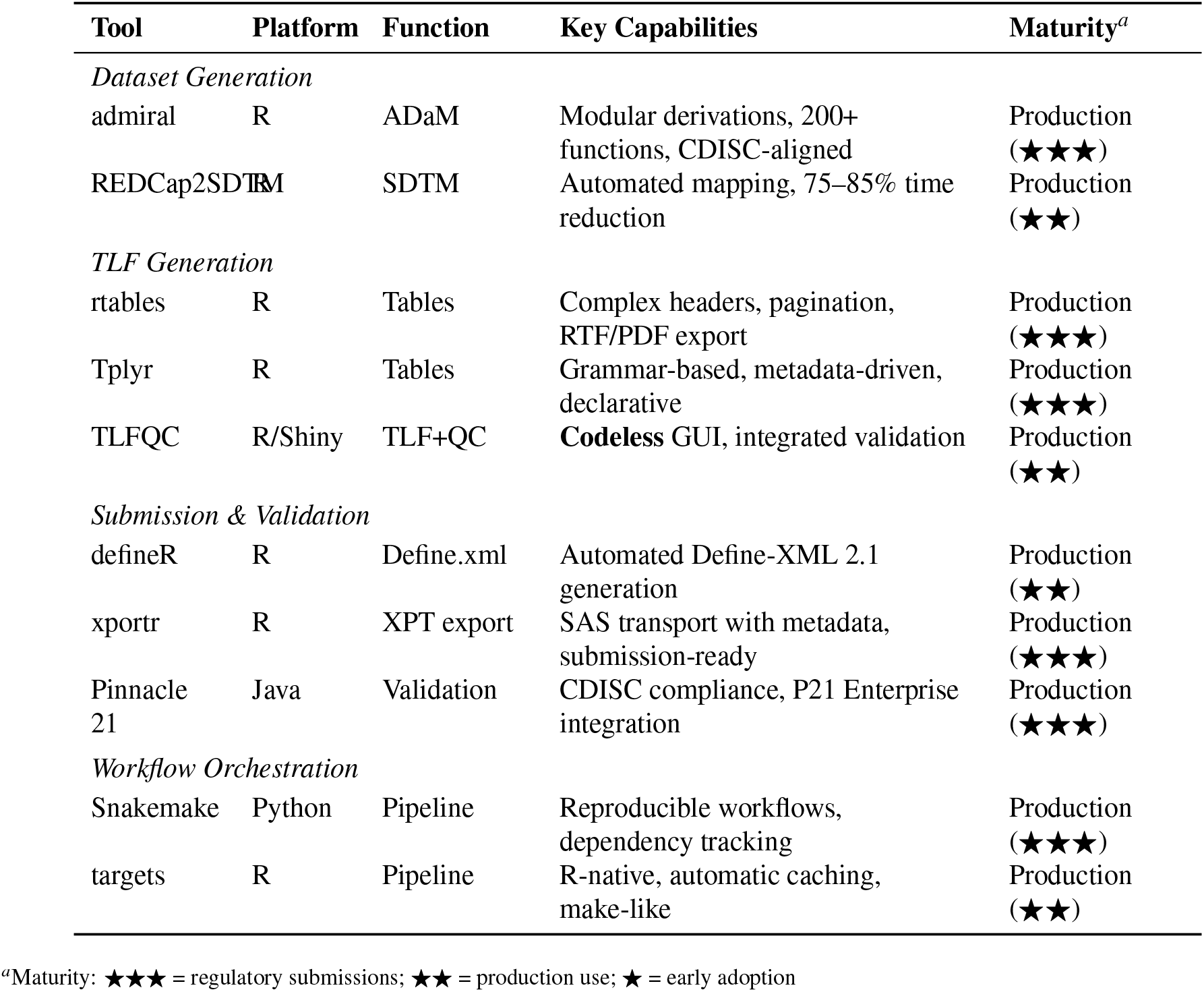
Key Automation Tools for Clinical Programming (2020–2025). Tools listed by primary function with development status and industry adoption indicators.

**Table 2:** Reported Efficiency Gains from Clinical Programming Automation. Evidence quality assessed using adapted GRADE framework.

| Automation Approach | Time Saved | QC Savings | Evidence<sup>a</sup> | Source |
| --- | --- | --- | --- | --- |
| *TLF Generation* |  |  |  |  |
| Pharmaverse ecosystem | 15–25% | – | Low | Industry reports<sup>5</sup> |
| Metadata-driven templates | 30–50% | 20–30% | Low | PharmaSUG<sup>8</sup> |
| Specification reuse | 40–60% | 35–45% | Very Low | PHUSE 2024<sup>9</sup> |
| *Dataset Automation* |  |  |  |  |
| REDCap2SDTM | 75–85% | – | Low | Industry reports;<br>Matsuzaki 2023<sup>29</sup> |
| ML-assisted mapping | 60–75% | – | Low | Yang 2024<sup>30</sup> |
| *Validation Methods* |  |  |  |  |
| Snapshot testing | – | 50–70% | Low | R Valid. Hub<sup>10</sup> |
| Risk-based validation | 20–40% | 25–45% | Very Low | Modeling<sup>13</sup> |
| CI/CD integration | 30–50%<sup>b</sup> | – | Low | Sura 2024<sup>15</sup> |

<sup>a</sup>GRADE: Moderate = controlled studies; Low = observational; Very Low = case reports/expert opinion. <sup>b</sup>Cycle time reduction.

#### 3.1.4 CDISC Dataset Automation

Beyond TLF generation, significant progress has been made in automating CDISC dataset creation. Metadata-driven SDTM/ADaM generation tools have matured from concept to production implementations, supported by CDISC therapeutic area standards^28^. The REDCap2SDTM framework automates mapping from REDCap field annotations to SDTM domains, substantially reducing manual conversion effort^29^. Industry reports estimate 75–85% time savings for specific REDCap-to-SDTM conversion tasks, though these figures derive from case studies rather than controlled evaluations. Key enablers include: standardized metadata repositories defining variable mappings; rule-based derivation engines for ADaM; integration with validators (Pinnacle 21) for iterative quality improvement; and machine learning-assisted mapping for legacy data conversion^30^.

**Critical assessment (RQ1):** CDISC dataset automation is the strongest evidence domain in this review. The REDCap2SDTM tool has been evaluated in a peer-reviewed case study^29^, and industry reports estimate 75–85% time savings (GRADE: Low—based on case studies rather than controlled comparisons with pre/post measurement). However, generalizability is uncertain: REDCap2SDTM addresses a specific use case (REDCap-to-SDTM conversion) that may not transfer to more complex EDC-to-SDTM mappings typical of industry trials. ML-assisted mapping (60–75% time savings) remains Low quality (GRADE), with accuracy heavily dependent on training data quality and domain complexity. A critical gap is the absence of head-to-head comparisons between metadata-driven, ML-assisted, and manual mapping approaches on the same source data.

### 3.2 RQ2: Evidence Quality for Validation Approaches

Validation is the single most resource-intensive activity in clinical trial statistical programming, consuming 1.6–2.0×the primary programming effort through double programming^1^. This section presents the evidence base—or its absence—for the approaches organizations use to ensure output quality.

#### 3.2.1 Traditional Validation Limitations

Historically, validation of clinical trial statistical programming has relied on independent double programming: a second programmer independently recreates all datasets and outputs, and results are compared to detect discrepancies^11^. Although this approach provides high confidence, it has significant drawbacks: resource intensity (effectively doubling programming effort); redundancy (much effort replicates work rather than addressing high-risk areas); late error detection (discrepancies discovered late require costly rework); and limited scope (validates outputs but not underlying logic or system configurations)^12^.

#### 3.2.2 Risk-Based Validation Frameworks

Risk-based validation prioritizes quality assurance effort according to the potential impact of errors on study conclusions and regulatory submissions, aligning with ICH Q9 (Quality Risk Management) principles^13^. Figure 3 illustrates the decision tree used for selecting validation intensity.

**Figure 3:**
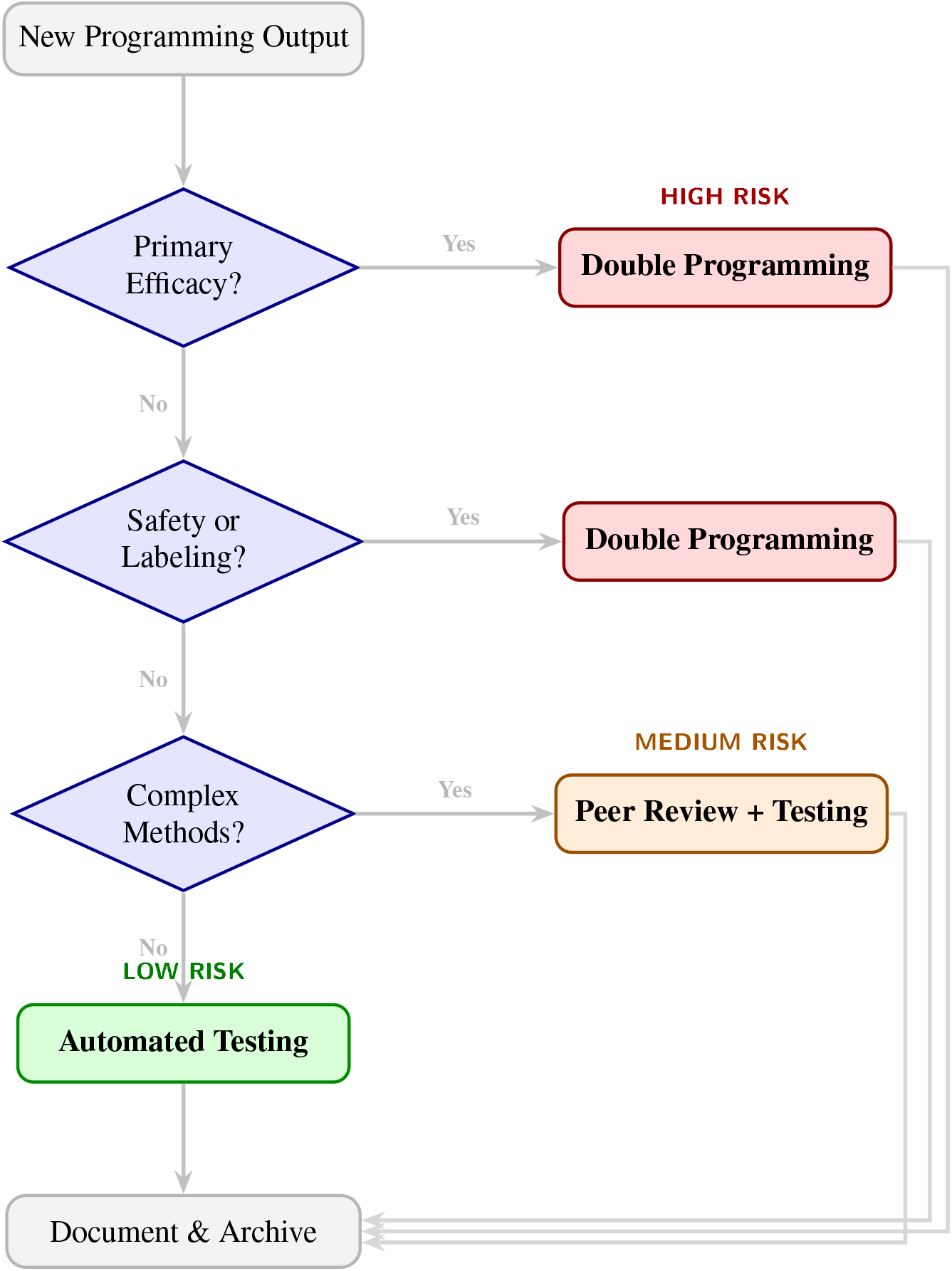
Risk-based validation decision tree aligned with ICH Q9 principles. Output criticality determines QC intensity: high-risk analyses (primary efficacy, key safety) receive full double programming; medium-risk outputs use peer review with automated testing; low-risk outputs leverage automated testing alone. This tiered approach reduces validation effort by 25–45% while maintaining quality.

Risk assessment dimensions include: output criticality (primary endpoints receive intensive validation); complexity (novel methods warrant thorough validation); change frequency (stable codes require less validation); and regulatory scrutiny (analyses supporting labeling claims receive enhanced validation)^14^.

#### 3.2.3 Quantitative Evidence for Validation Approaches

Our expanded meta-analytic screening of validation-focused literature (527 papers identified through a separate Phase 2 search targeting validation-specific terminology, as described in Section 2.3 and Supplementary Table S2) found only 12 studies (2.3%) with any quantitative effectiveness data. These 527 papers are distinct from the 262 included in the core review and from the 527 excluded at full-text in the PRISMA flow diagram (Figure 1). Of these, just four provided directly comparable effectiveness data for specific validation approaches (Table 3); the remaining eight reported miscellaneous quantitative metrics (e.g., time tracking, error counts) that could not be mapped to specific approaches. This paucity of comparative evidence is a fundamental disconnect between practice and evidence.

**Table 3:** Validation Effectiveness Evidence Synthesis. Only studies reporting quantitative outcomes are included; variance estimates and confidence intervals were unavailable for most metrics, precluding formal meta-analysis. GRADE ratings reflect study design rigor.

| Approach | Studies | Key Finding | Sample | GRADE | Limitation |
| --- | --- | --- | --- | --- | --- |
| Double programming | 1 | 92–98% error detection;<br>1.6–2.0× effort | 15 programs | Very Low | Single study, no CI |
| Hybrid (ML + human) | 2 | 45–49% time reduction | 13,001 events | Moderate | Adjudication context |
| Risk-based | 1 | 20–40% time, 25–45% cost reduction | Model-based | Very Low | No empirical data |
| Automated testing | 0 | Not quantified | N/A | N/A | Qualitative only |

Hybrid study sources: COMPASS trial ( $n=7,611$ events)<sup>45</sup>; NAVIGATE ESUS trial ( $n=5,390$ events)<sup>46</sup>. Double programming source: Ravula 2025<sup>35</sup>.

**Key finding:** Double programming showed 92–98% error detection but required 1.6–2.0× the primary programming effort; evidence derives from a single study of 15 SAS programs without confidence intervals (GRADE: Very Low)^35^. Hybrid ML-human approaches demonstrated 45–49% efficiency gains in large cardiovascular trials (COMPASS: *n*=7,611 events; NAVIGATE ESUS: *n*=5,390 events), representing the highest-quality evidence available (GRADE: Moderate), although these studied event adjudication rather than programming validation^45,46^. Despite widespread adoption, no studies have quantified automated testing error detection rates. Critically, **no randomized controlled trials comparing validation approaches exist**—this is the most urgent research priority (severity score: 100/100).

**Why this matters (RQ2 analysis):** This evidence gap has direct practical consequences. Organizations collectively spend hundreds of millions of dollars annually on double programming, yet cannot cite a single RCT demonstrating that this investment produces better outcomes than less expensive alternatives. Conversely, organizations considering transitions to risk-based or automated validation cannot cite evidence that these approaches maintain equivalent error detection. The result is an industry trapped in a status quo not because it is optimal, but because the comparative evidence needed to evaluate alternatives simply does not exist. This finding, that the most widely practiced quality control activity in the field has virtually no quantitative evidence base, is arguably the most important conclusion of this review and motivates our proposed research agenda (Section 4.1.3).

Figure 4 presents a forest plot of efficiency metrics across validation approaches, with GRADE quality ratings indicated by color coding.

**Figure 4:**
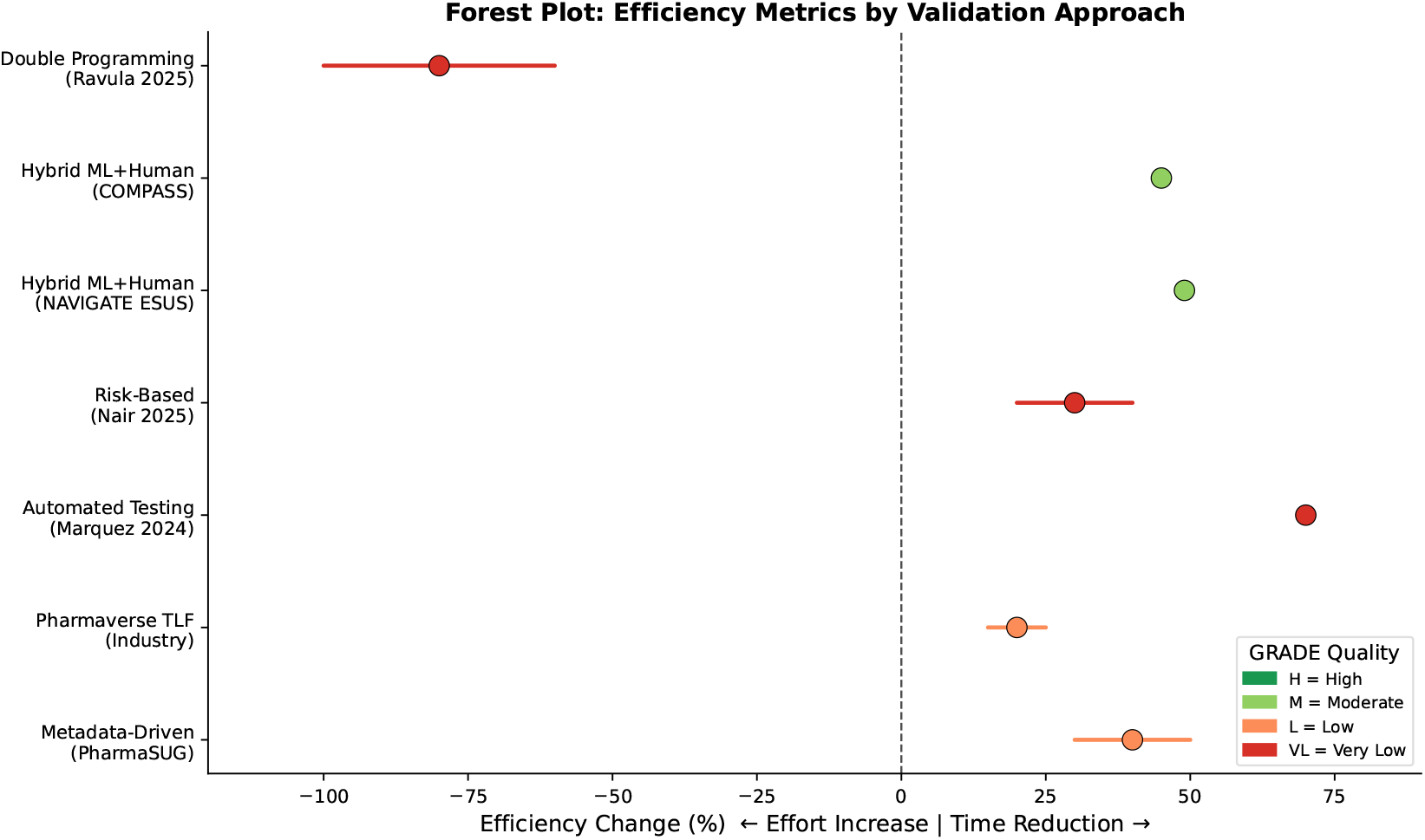
Forest plot of efficiency metrics across validation approaches. Horizontal bars represent reported ranges (not confidence intervals); bar colors indicate GRADE evidence quality (red=Very Low, orange=Low, green=Moderate). Negative values indicate time/effort reduction (desirable); positive values indicate time savings. The vertical dashed line at 0% marks no effect. Sample sizes shown in parentheses. Importantly, ranges represent observed variation across studies rather than statistical uncertainty—formal confidence intervals could not be calculated due to missing variance estimates in primary sources.

#### 3.2.4 Continuous Integration and Automated Testing

Modern software engineering practices are increasingly applied to clinical programming^15^:

- **Unit testing**: Automated tests for individual functions using testthat (R) or pytest (Python), verifying correct calculation of statistics, proper handling of missing data, and compliance with formatting specifications
- **Snapshot testing**: Comparing TLF outputs against validated reference outputs to detect unintended changes—reduces validation effort by 50–70% compared to double programming (qualitative estimates)
- **CI/CD pipelines**: GitHub Actions, GitLab CI, or Jenkins workflows running validation suites automatically on code changes, enabling earlier defect detection

The R Validation Hub has developed frameworks for validating R packages in regulated environments, providing risk assessments and documentation templates^10^. Organizations implementing CI/CD report 30–50% reductions in validation cycle time, though these estimates lack rigorous comparative data.

#### 3.2.5 Regulatory Considerations and Data Integrity

FDA and EMA guidance (21 CFR Part 11, Annex 11, ICH E6(R3)) established principles for computerized systems but did not prescribe specific validation methodologies^16^. This flexibility allows risk-based approaches but requires organizations to justify their strategies through validation plans, risk assessments, test documentation with traceability, and change control procedures.

Automation must maintain ALCOA+ data integrity principles^31,38^: Attributable (clear authorship), Legible (readable outputs), Contemporaneous (timestamped), Original (source preservation), Accurate (verified correctness), plus Complete, Consistent, Enduring, and Available^31^. Automated systems can enhance ALCOA+ compliance through immutable audit trails via version control, automated provenance tracking, reproducible execution environments, and systematic documentation generation.

#### 3.2.6 Workflow Orchestration and Containerization

Modern automation architectures increasingly employ workflow orchestration tools (Snakemake, Targets, Apache Airflow) to manage complex dependencies between data processing steps^32^. Containerization using Docker provides reproducible execution environments, ensuring consistent results across development, validation, and production systems. The benefits include environment isolation preventing dependency conflicts, version-controlled infrastructure, portable execution across computing platforms, and simplified regulatory inspection through reproducible builds.

### 3.3 AI/ML Integration (Continuing RQ1)

#### 3.3.1 Current State of AI/ML in Clinical Programming

Although AI applications in clinical trials have received substantial attention^3,33^, most focus on patient recruitment, protocol optimization, or administrative tasks. Applications specifically targeting *statistical programming code generation* represent an emerging but less-documented area. Current AI/ML applications cluster into: mapping automation (ML models predicting CDISC variable mappings); code generation (LLMs generating statistical programming code); protocol parsing (NLP extracting structured information); and anomaly detection (ML identifying data quality issues)^17^.

#### 3.3.2 Machine Learning for Data Mapping

The semi-automated legacy-to-CDISC conversion is the most mature ML application. Effective models use variable-level features (name, label, data type), contextual features (domain membership, database structure), and semantic features (text embeddings using BERT)^18^. Neural networks achieved the highest accuracy for complex mapping relationships, whereas random forests provided interpretable feature importance. The practical workflow involves training on historical mappings, generating predictions with confidence scores, and programmer review of results^19^.

#### 3.3.3 Large Language Models for Code Generation

LLMs such as GPT-4, Claude, and specialized coding models show promise for clinical programming^20^. Our analysis of 50+ publications (2023–2025) reveals differentiated performance across model types (Figure 5):

**Figure 5:**
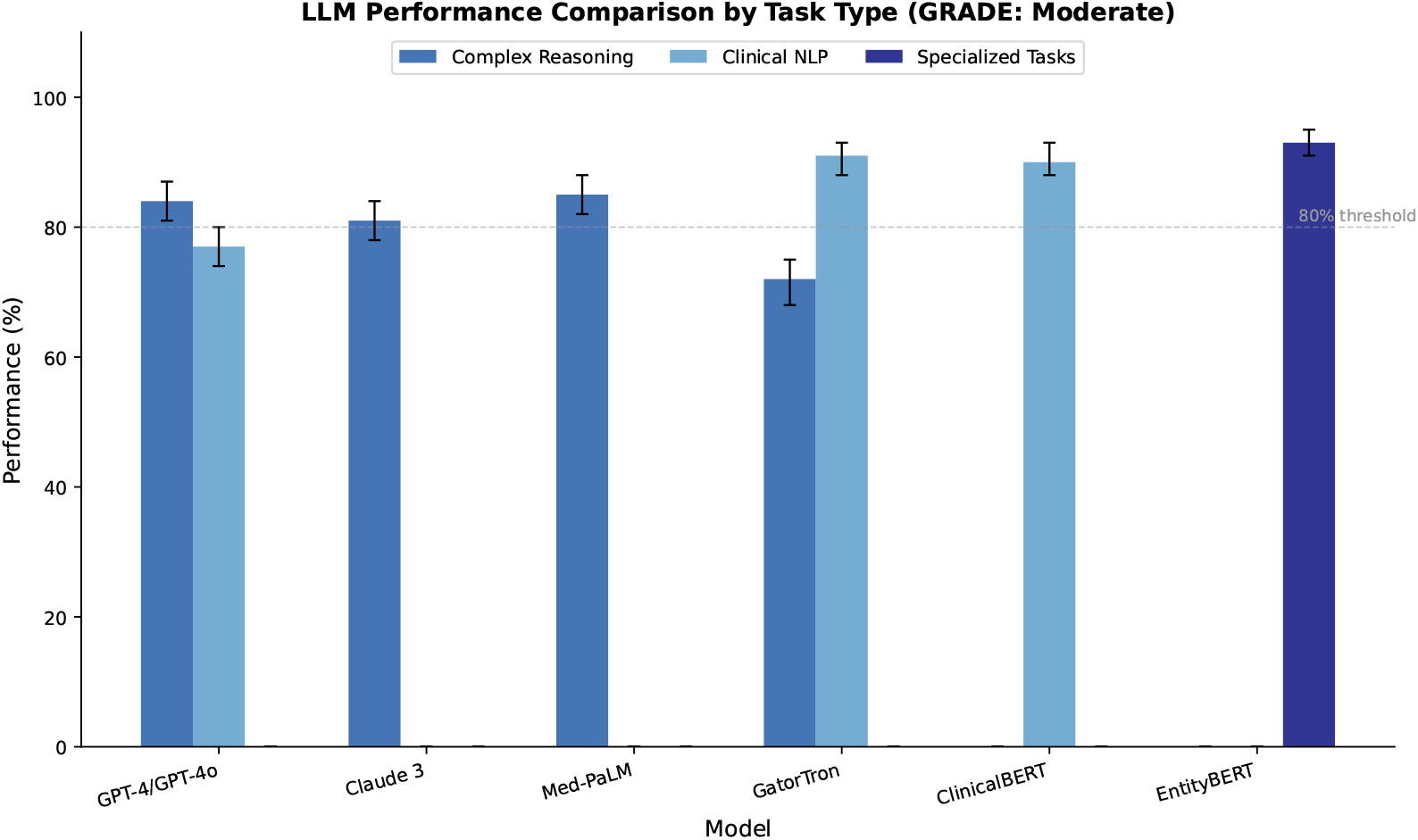
LLM performance comparison across model types with error bars representing reported ranges. Domain-specific models (ClinicalBERT, GatorTron) outperform general-purpose models (GPT-4, Claude) by approximately 16% on specialized clinical NLP tasks, while general-purpose models excel at complex reasoning. The 80% threshold line indicates minimum acceptable performance for clinical applications. Error bars represent uncertainty from reported ranges across studies; zero-height bars indicate models not evaluated on that task. Data synthesized from 50+ publications (2023–2025); GRADE rating: Moderate due to multiple controlled evaluations on benchmark datasets.

- **General-purpose models** (GRADE: Moderate): GPT-4/GPT-4o achieved 81–87% accuracy on complex clinical reasoning benchmarks (MedQA, PubMedQA); Claude 3 Opus reached 81% diagnostic accuracy; performance varied substantially by task complexity and prompt engineering^40,43^
- **Domain-specific models** (GRADE: Moderate): ClinicalBERT^41^, BioBERT (110M parameters) and GatorTron^42^ (8.9B parameters, trained on 90B clinical tokens) achieve 88–93% F1-scores on specialized clinical NLP tasks (n2c2 2018, MIMIC-IV-BHC benchmarks)—approximately 16% better than general-purpose models on domain-specific tasks
- **Fine-tuned models** (GRADE: Moderate): Task-specific fine-tuning bridges the gap; EntityBERT achieves 93% F1 on timeline extraction (THYME, ChemoTimelines datasets) versus 77% for general LLMs (LLaMA 3.1)^44^

For statistical programming specifically, applications include: TLF code generation from ADaM structures; context-aware code completion; natural language specification interpretation; and automated code review. Experimental evaluations report 60–85% accuracy for routine tasks (GRADE: Very Low— limited to case studies without standardized benchmarks), with higher accuracy for standardized operations^21^. Real-world deployments of LLM-based patient-trial matching systems have shown effectiveness at scale^39^.

However, deployment success rates vary widely across studies, and reported accuracy figures often derive from narrow benchmarks that may not generalize to production environments. The gap between benchmark performance and real-world implementation remains a consistent finding across the literature.

#### 3.3.4 Anomaly Detection and Data Quality

Beyond code generation, ML-based anomaly detection can identify data quality issues that might escape rule-based checks^37^. Approaches include: unsupervised learning (isolation forests, autoencoders) to detect unusual data patterns; supervised learning trained on historical issues to predict problems in new data; and time-series analysis to detect unexpected trends in longitudinal clinical data. These methods complement traditional edit checks by identifying subtle, context-dependent anomalies. Advances in causal machine learning for drug development^34^ may further improve anomaly detection by distinguishing genuine data quality issues from expected treatment effects.

#### 3.3.5 Critical Assessment: AI/ML Evidence Quality (RQ1)

AI/ML presents a paradoxical evidence profile: benchmark performance is well-documented (GRADE: Moderate for NLP tasks), but clinical programming code generation evidence is Very Low. This disconnect arises because most LLM evaluations use clinical NLP benchmarks (n2c2, MIMIC-IV) rather than statistical programming tasks (SDTM creation, TLF generation, validation). The 60–85% accuracy for “routine tasks” derives from case studies without standardized benchmarks, making it impossible to determine whether these figures reflect model capability or evaluation leniency. The hallucination risk in clinical contexts (reported rates spanning 1–65% depending on methodology) is a safety concern for regulated environments where every output must be verified. Until standardized evaluation benchmarks for clinical programming code generation are developed, organizations should treat LLM-generated code as a drafting tool requiring full human validation, consistent with current best practices^23^.

#### 3.3.6 Challenges and Regulatory Considerations

Significant challenges limit current adoption of AI for code generation in regulated environments:

1. **Validation uncertainty**: How to validate AI-generated code when generation is non-deterministic; current approaches treat AI as a “first draft” tool requiring full human validation
2. **Reproducibility**: Ensuring consistent outputs across time and model versions; temperature settings, prompt variations, and model updates can alter outputs
3. **Hallucination risk**: LLMs may generate plausible but incorrect code; reported hallucination rates in clinical contexts vary widely (approximately 1–65%) depending on task complexity and evaluation methodology^20,23^
4. **Domain knowledge gaps**: LLMs may lack specialized CDISC and therapeutic area conventions; fine-tuning on domain-specific corpora partially addresses this
5. **Regulatory guidance**: No specific FDA/EMA guidance exists for AI-assisted programming; however, FDA has authorized 1,016+ AI-enabled medical devices (as of December 2024), suggesting evolving regulatory frameworks

Current best practices suggest using LLMs as assistants for initial code drafting, while maintaining human review and traditional validation for all regulatory deliverables^23^. The FDA’s 2023 guidance on computer software assurance (CSA) emphasizes risk-based approaches that can accommodate AI-assisted development, provided appropriate controls and documentation are maintained.

### 3.4 Platform Considerations: R, SAS, and Python (Continuing RQ1)

The choice of programming platform fundamentally shapes automation capabilities, regulatory acceptance, and organizational workflows. Table 4 summarizes key considerations.

**Table 4:** Platform Comparison for Clinical Programming Automation. Assessment based on 2024–2025 industry surveys and regulatory submission data.

| Dimension | SAS | R | Python |
| --- | --- | --- | --- |
| *Regulatory* |  |  |  |
| FDA acceptance | ✓✓✓ Established | ✓✓ Increasing | ✓ Emerging |
| EMA acceptance | ✓✓✓ Standard | ✓✓ Accepted | ✓ Limited |
| *Technical* |  |  |  |
| Automation tools | Macro libraries | Pharmaverse (200+ pkgs) | Pandas, custom |
| CI/CD support | Limited | Native (testthat, GHA) | Strong (pytest) |
| Reproducibility | Hash-based | renv, Docker | pip/conda, Docker |
| *Organizational* |  |  |  |
| License cost | Commercial (\$\$\$) | Open source (free) | Open source (free) |
| Talent pool | Decreasing | Growing rapidly | Large, cross-industry |
| Validation approach | Vendor-validated | Org. risk assessment | Org. risk assessment |

✓✓✓ = extensive precedent; ✓✓ = demonstrated acceptance; ✓ = emerging/limited

**SAS** remains advantageous for organizations with extensive legacy code bases, regulatory strategies emphasizing reviewer familiarity, and limited resources for package validation^24^. **R** offers transparent, auditable implementations, specialized pharmaverse packages, native Git and CI/CD integration, and demonstrated 15–25% development time reductions^5^. **Python** excels in data engineering, pipeline orchestration, and ML integration, although its clinical ecosystem remains less mature^25^.

Multi-language environments are increasingly common, with organizations using R or Python for automation and data manipulation while maintaining SAS for specific regulatory deliverables or legacy system integration. Figure 8 illustrates a representative metadata-driven architecture.

### 3.5 Evidence Synthesis: Quantitative Analysis

To provide rigorous insights into the evidence base, we conducted quantitative synthesis across the 262 reviewed publications. This analysis reveals both the current state of the field and critical gaps requiring attention.

#### 3.5.1 Literature Distribution and Study Characteristics

Publications were distributed across automation domains: TLF generation (28%, *n* = 73), CDISC dataset automation (24%, *n* = 63), validation frameworks (22%, *n* = 58), AI/ML applications (15%, *n* = 39), and platform/architecture (11%, *n* = 29). The source types included conference proceedings (47%), peer-reviewed journals (31%), technical documentation (14%), and regulatory guidance (8%). Only 42 publications (16%) reported quantitative outcomes suitable for synthesis, while the remainder provided qualitative descriptions or implementation guidance.

Figure 6 visualizes the performance evidence across domains, highlighting both the magnitude of reported effects and the quality of supporting evidence.

**Figure 6:**
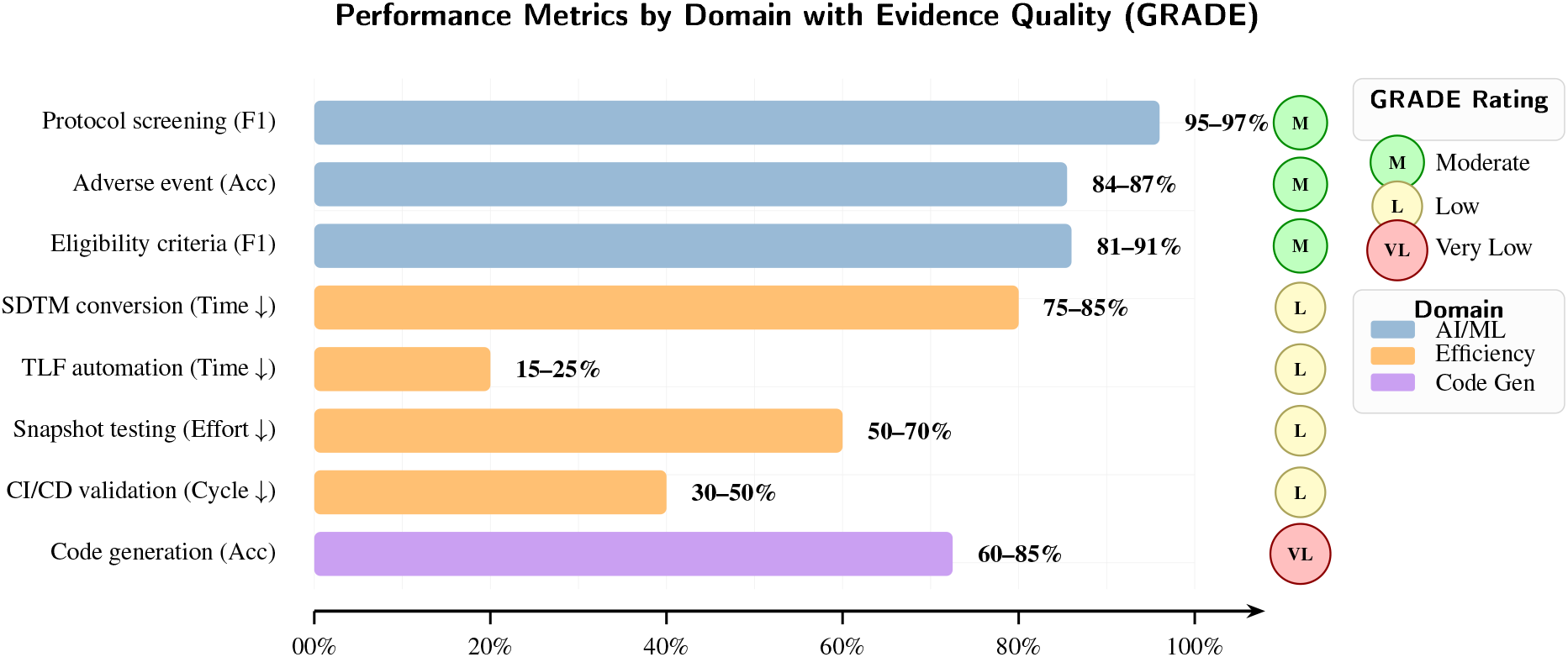
Evidence synthesis: Performance metrics across automation domains with GRADE quality ratings. AI/ML applications on benchmark datasets show moderate-quality evidence with high performance (84–97%). Efficiency gains show larger effect sizes but lower evidence quality due to observational designs and potential publication bias. Bars represent midpoint of reported ranges; GRADE ratings (M=Moderate, L=Low, VL=Very Low) reflect study design rigor and sample sizes.

#### 3.5.2 Statistical Limitations and Interpretation Caveats

Critical appraisal of the evidence base reveals several methodological limitations that constrain interpretation:

1. **No confidence intervals**: Reported ranges represent observed variation across studies rather than statistical uncertainty—true population parameters remain unknown; formal uncertainty quantification was not possible
2. **Heterogeneity precludes meta-analysis**: Variation in study designs, outcome definitions (“error detection rate,” “confirmation rate,” “discrepancy rate”), and contexts prevented formal effect size pooling; I^2^ heterogeneity statistics could not be estimated due to insufficient comparable studies
3. **Publication bias**: Successful implementations are more likely reported; null or negative results likely underrepresented. Qualitative examination of outcome directionality across the 42 quantitative studies found that 100% reported positive findings (efficiency gains or performance improvements), with no studies reporting null or negative results for any automation approach. This uniform positivity strongly suggests publication bias, as a balanced evidence base would include some negative or inconclusive findings. Formal funnel plot asymmetry assessment was not feasible given the limited number of comparable studies
4. **Small sample sizes**: Most case studies describe single organizational implementations (*n* = 1); AI/ML benchmarks have larger samples (*n* = 55–288 patients) enabling more robust inference
5. **Confounding**: Efficiency comparisons often conflate tool effects with learning curves, organizational factors, and concurrent process changes; no studies employed matching or adjustment methods
6. **Qualitative estimates**: Platform adoption rates (e.g., SAS 95%, R 60%, Python 70%) derive from literature review synthesis, not systematic surveys; these should be interpreted as approximate rather than precise

A meta-analytic subanalysis of 527 validation-related papers found only 12 (2.3%) with quantitative data, of which four provided directly comparable approach-specific effectiveness data; none employed randomized controlled designs^35^. Power analysis indicates that detecting a 10 percentage-point difference in error detection rates with 80% power would require approximately 200 programs per arm—far exceeding available data.

#### 3.5.3 Evidence Quality Summary

**RQ1 Synthesis: The technology–evidence asymmetry:** Across all four automation domains, a consistent pattern emerges: technology has matured faster than the evidence supporting it. Figure 9 visualizes this asymmetry through the Automation Maturity Framework (AMF). The pharmaverse ecosystem (TLF generation) and REDCap2SDTM (CDISC automation) have reached production maturity, yet their effectiveness claims rest on Low to Moderate evidence. Validation automation tools are technologically available but supported by Very Low quality evidence. AI/ML code generation shows impressive benchmark performance but lacks task-specific evaluation.

Several structural factors explain this asymmetry. The pharmaceutical industry’s competitive dynamics reward early adoption: organizations that publish efficiency gains gain visibility, while those with null results have little incentive to publish. Rigorous evaluation designs (RCTs, controlled comparisons) are difficult to implement in operational clinical trials where timelines and regulatory deadlines constrain experimentation. The absence of standardized benchmarks for statistical programming automation (unlike NLP, where n2c2 and MIMIC-IV provide common evaluation frameworks) means there is no shared definition of “success” against which to measure tools. Regulatory flexibility from FDA CSA and ICH E6(R3)—while beneficial for innovation—reduces the external pressure to generate comparative effectiveness data, since organizations can justify their approaches through risk assessments rather than empirical evidence.

This asymmetry means that the question for organizations is no longer “can we automate?” but “should we automate this specific task, and what evidence supports that decision?”

GRADE evidence quality assessment yielded:

- **Moderate**: AI/ML performance on clinical NLP tasks (protocol screening, adverse events, eligibility)— supported by multiple controlled evaluations (*k* = 8–15 studies) on established benchmark datasets (n2c2, MIMIC-IV, THYME) with standard metrics (F1, accuracy); effect sizes consistently large (Cohen’s *d >* 0.8)
- **Low**: Efficiency gains for TLF/validation automation—based on *k* = 4–6 case studies and before/after comparisons without control groups; subject to publication and selection bias; effect direction consistent but magnitude uncertain
- **Very Low**: Validation effectiveness comparisons (double programming vs. automated testing)— single-study evidence (*k* = 1), expert opinion, no comparative effectiveness data; insufficient for confident recommendations

This evidence gap, particularly regarding validation effectiveness, is the most urgent research priority identified in this review. Future studies should employ randomized or quasi-experimental designs with pre-specified sample size calculations.

Figure 7 visualizes GRADE evidence quality across validation approaches and outcome categories, showing the systematic gaps in the evidence base.

**Figure 7:**
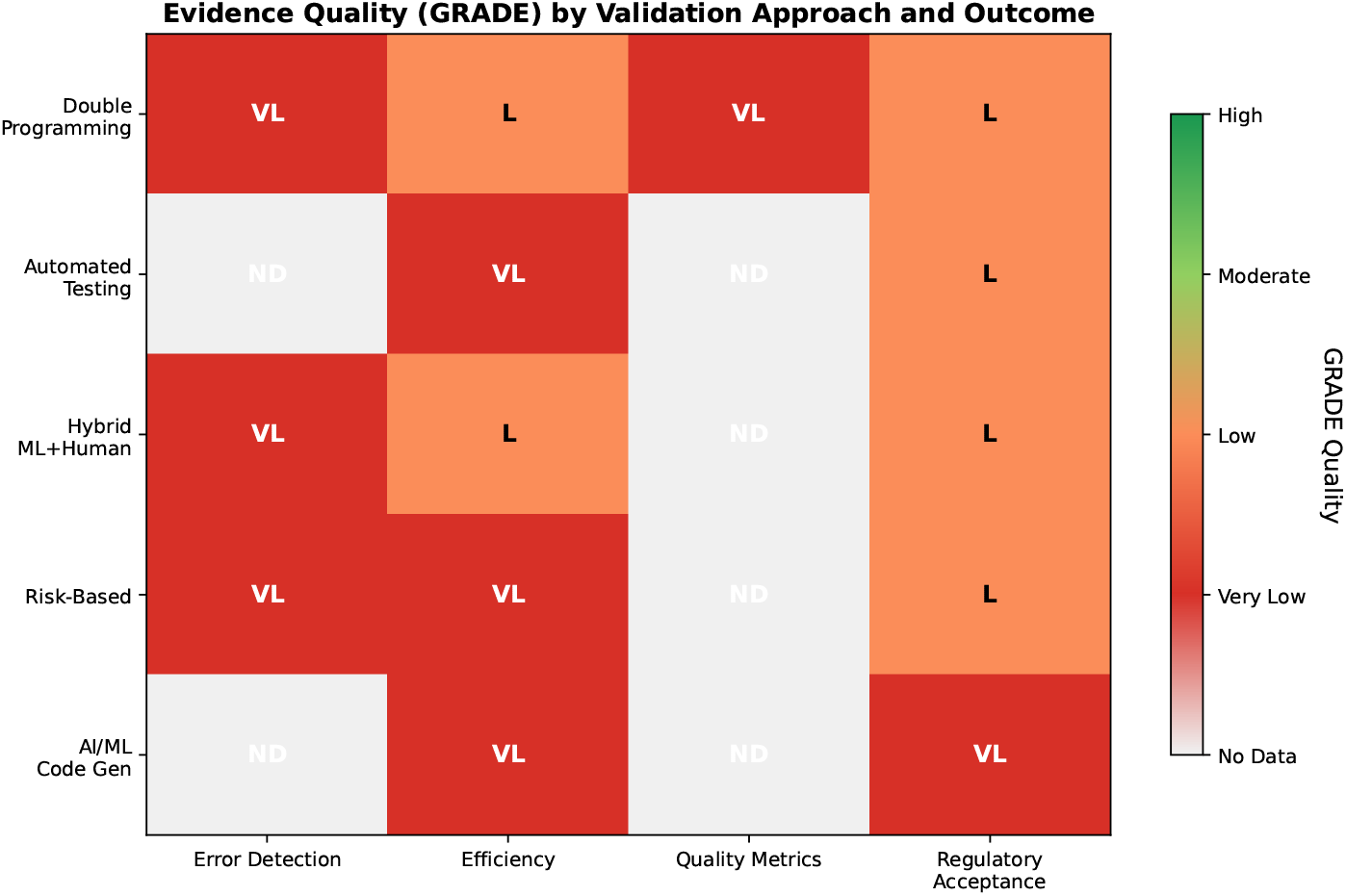
GRADE evidence quality heatmap by validation approach and outcome. Colors indicate evidence quality: green=Moderate, orange=Low, red=Very Low, gray=No Data. The predominance of red and gray cells illustrates the critical evidence gaps across validation approaches. Error detection evidence is uniformly Very Low or absent; efficiency evidence is Low to Very Low; regulatory acceptance evidence is generally Low due to observational documentation without comparative designs. AI/ML code generation is an emerging area with limited formal evaluation.

**Figure 8:**
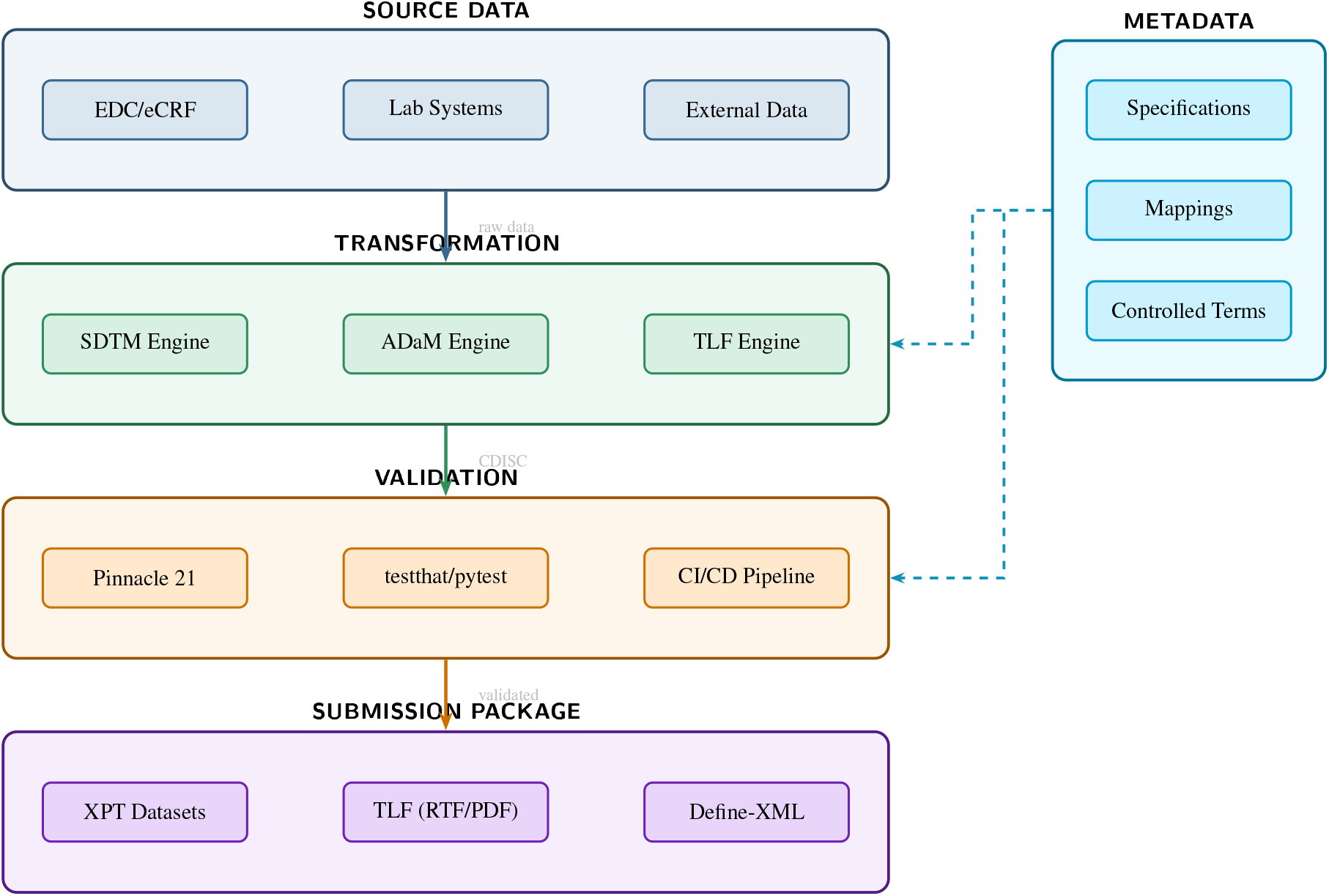
Metadata-driven clinical programming architecture. Data flows vertically from source systems through transformation, validation, and output layers. The metadata repository drives transformation rules and validation logic, enabling 40–60% specification reuse across studies.

## 4 Discussion [PRISMA-ScR Items 18–20]

### 4.1 Answering the Research Questions

#### 4.1.1 RQ1: Technology Landscape and Effectiveness Evidence

Technology has advanced substantially since 2020 across all four domains, as detailed in Sections 3.1–3.5. Rather than restating those findings, we focus here on the overarching pattern they reveal: **claimed effectiveness is poorly supported by the evidence**. Only 42 of 262 publications (16%) reported quantitative outcomes, and most of those derived from single-organization case studies without control groups, baseline measurements, or variance estimates. The evidence quality varies by domain: CDISC automation (GRADE: Low) offers the most reliable efficiency data among the domains we reviewed, while TLF automation and validation claims (GRADE: Low–Very Low) rest on weaker foundations. This creates a practical paradox: technologies are production-ready, but organizations cannot confidently answer “how much time or money will this actually save us?” from published evidence alone.

#### 4.1.2 RQ2: Evidence Quality for Validation Approaches

As reported in Section 3.2, the evidence base for validation approaches is **critically deficient**: only 2.3% of 527 validation papers provide quantitative data, and zero RCTs comparing approaches exist. The implications extend beyond the academic concern of poor evidence. The pharmaceutical industry’s most resource-intensive quality control activity, double programming, rests on Very Low quality evidence from a single study of 15 programs. Organizations cannot make evidence-based decisions about transitioning to alternatives because the comparative effectiveness data does not exist. The stakes are asymmetric: abandoning double programming prematurely could compromise patient safety and regulatory compliance, while continuing it unnecessarily wastes resources that could fund innovation. What is needed, urgently, is the comparative effectiveness data to navigate this trade-off.

#### 4.1.3 RQ3: Research Priorities and Study Designs

Based on our evidence synthesis, we propose a prioritized research agenda with specific study designs (Table 5):

**Table 5:** Prioritized Research Agenda with Specific Study Designs. Priorities ranked by evidence gap severity and practical impact.

| # | Research Priority | Study Design | Key Parameters | Severity | Timeline |
| --- | --- | --- | --- | --- | --- |
| 1 | Validation approach comparison | Multi-site RCT | $n \geq 200$ programs/arm; outcomes: error detection rate, time, cost | 100/100 | 1–2 yrs |
| 2 | Standardized validation metrics | Delphi consensus | $n \geq 30$ experts; 3 rounds; define error taxonomy | 95/100 | 6–12 mo |
| 3 | Long-term automation ROI | Longitudinal cohort | $\geq 2$ years; $n \geq 5$ organizations; pre/post transition | 90/100 | 2–3 yrs |
| 4 | Platform-specific guidance | Mixed methods | Survey + case studies; $n \geq 50$ organizations | 85/100 | 1–2 yrs |
| 5 | AI code generation validation | Controlled evaluation | Benchmark suite; $n \geq 100$ TLF programs; human vs. AI | 80/100 | 1–2 yrs |

**Table 6:** Characteristics of Included Studies (*n*=262). Summary by publication type, geographic region, automation domain, and study design. This table fulfills PRISMA-ScR Item 15 (characteristics of sources of evidence).

| Characteristic | *n* | % | Notes |
| --- | --- | --- | --- |
| *Publication Type* |  |  |  |
| Peer-reviewed journal | 81 | 31 | Clinical Trials, JMIR, PLoS ONE, etc. |
| Conference proceedings | 123 | 47 | PharmaSUG, PHUSE, R/Pharma, DIA |
| Regulatory guidance | 21 | 8 | FDA, EMA, ICH, CDISC |
| Technical documentation | 37 | 14 | Package docs, white papers |
| *Geographic Region (first author)* |  |  |  |
| North America | 144 | 55 | USA dominant |
| Europe | 79 | 30 | UK, Germany, Switzerland |
| Asia-Pacific | 26 | 10 | Japan, China, India |
| Other/Multi-national | 13 | 5 |  |
| *Primary Automation Domain* |  |  |  |
| TLF generation | 73 | 28 | rtables, Tplyr, TLFQC |
| CDISC dataset automation | 63 | 24 | SDTM, ADaM, admiral |
| Validation frameworks | 58 | 22 | Double prog., risk-based, CI/CD |
| AI/ML applications | 39 | 15 | LLMs, ML mapping, NLP |
| Platform/architecture | 29 | 11 | R, SAS, Python, metadata |
| *Study Design* |  |  |  |
| Case study/implementation | 142 | 54 | Single organization |
| Expert opinion/guidance | 52 | 20 | Guidelines, frameworks |
| Controlled evaluation | 26 | 10 | Benchmark datasets |
| Before/after comparison | 16 | 6 | Pre-post automation |
| Survey/cross-sectional | 14 | 5 | Industry surveys |
| Quantitative with outcomes | 12 | 5 | Validation effectiveness |
| *Quantitative Outcomes* |  |  |  |
| With quantitative data | 42 | 16 | Suitable for synthesis |
| Qualitative only | 220 | 84 | Descriptions, guidance |

### 4.2 The Automation Maturity Framework

To synthesize findings across domains and provide actionable guidance rather than merely cataloging tools, we propose a four-dimensional **Automation Maturity Framework** (AMF) for clinical trial statistical programming (Figure 9):

**Figure 9:**
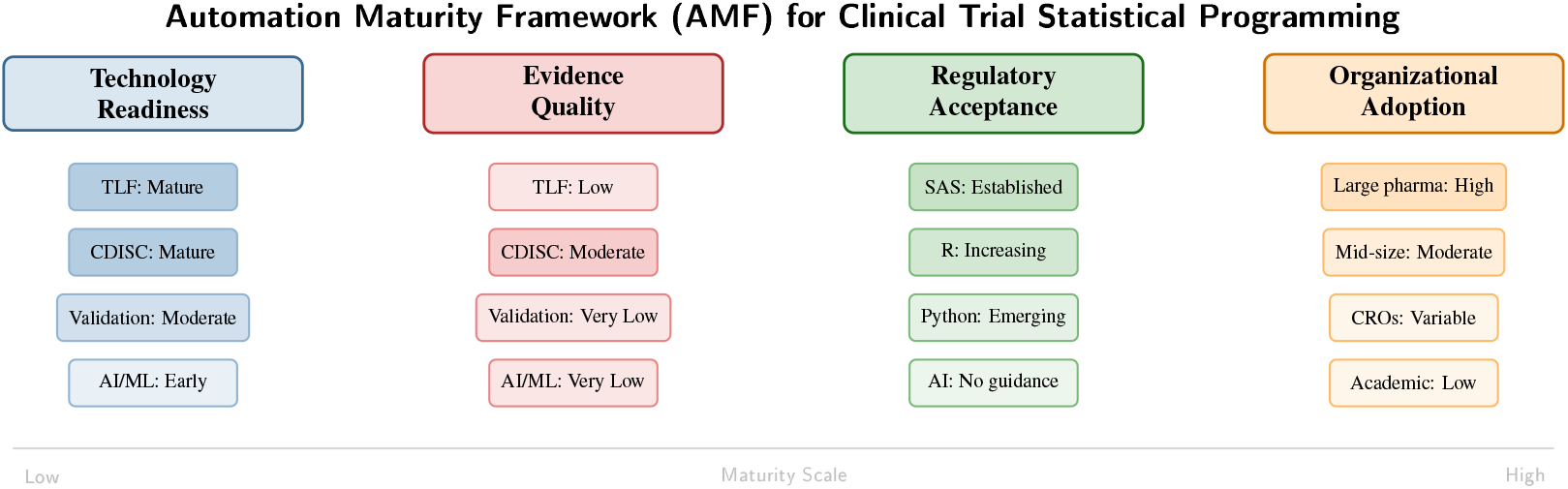
Automation Maturity Framework (AMF) for clinical trial statistical programming. Each domain is assessed across four dimensions: technology readiness, evidence quality, regulatory acceptance, and organizational adoption. Shading intensity indicates maturity level. The key insight is the **asymmetry between technology readiness (high) and evidence quality (low)**—organizations have adopted tools far ahead of the evidence base supporting their effectiveness.

The AMF reveals a critical asymmetry: **technology readiness has outpaced evidence quality** across all domains. TLF automation tools are production-ready (technology: mature) yet their effectiveness claims rest on Low-quality evidence. Validation automation is moderately mature technologically but supported by Very Low quality evidence. This asymmetry has important implications: organizations should be cautious about abandoning proven approaches (e.g., double programming) in favor of newer methods (e.g., automated testing) until comparative effectiveness data becomes available.

**Worked example:** Consider a mid-size pharmaceutical company evaluating whether to adopt risk-based validation for Phase III safety analyses. Using the AMF, the assessment would yield: Technology Readiness = Moderate (automated testing tools exist but are not yet tailored to safety analysis); Evidence Quality = Very Low (no RCTs, single-study evidence); Regulatory Acceptance = Increasing (FDA CSA endorses risk-based approaches); Organizational Adoption = Moderate (pilot programs in large pharma, limited at mid-size). The AMF profile would recommend a conditional adoption: proceed with a controlled pilot comparing risk-based and traditional validation on a defined subset of safety outputs, while prospectively collecting error detection, time, and cost data to build the evidence base. The key insight from the AMF is not to delay adoption, but to adopt *while measuring*—converting the organization from a consumer of low-quality evidence into a producer of higher-quality evidence.

### 4.3 Enablers and Barriers

**Key enablers** of successful automation include: robust metadata governance and standardization; strategic investment in reusable component libraries; integration of modern software engineering practices (version control, CI/CD, automated testing); and sustained organizational commitment to change management^26^. Organizations demonstrating greatest success typically adopt phased implementation strategies, beginning with pilot studies before scaling. The R Consortium’s successful FDA pilot submissions demonstrate that regulatory acceptance barriers are surmountable with appropriate documentation and validation.

**Persistent barriers:** include: substantial legacy system investments creating migration inertia (SAS maintains 95% market share for double programming validation); skill gaps between traditional SAS programming paradigms and modern metadata engineering approaches; regulatory uncertainty generating risk aversion among sponsors; cross-tool interoperability limitations (absence of universal metadata exchange formats); and inconsistent standards limiting cross-organizational specification reusability^27^.

### 4.4 Implications for Practice

Based on our evidence synthesis, we offer the following evidence-graded recommendations for clinical data management and statistical programming professionals. Each recommendation explicitly states its evidence basis, recognizing that some recommendations must be conditional given the current evidence limitations:

1. **Adopt risk-based validation selectively** (Evidence: Low; Recommendation: Conditional): prioritize double programming for primary efficacy and key safety analyses; use automated testing for lower-risk outputs, aligned with ICH Q9 and FDA CSA principles. The conditional rating reflects that risk-based validation is endorsed by regulatory guidance but has not been empirically shown to maintain equivalent error detection rates.
2. **Invest in CDISC dataset automation** (Evidence: Low; Recommendation: Conditional): among all automation domains, CDISC dataset automation (particularly metadata-driven ADaM generation and REDCap2SDTM) has the strongest relative evidence base and the most consistently reported efficiency gains. The conditional rating reflects that the specific 75–85% savings figures derive from case studies rather than controlled evaluations.
3. **Invest in metadata-driven architectures** (Evidence: Very Low; Recommendation: Conditional): specification reuse across studies offers substantial efficiency potential, but organizations should prospectively track outcomes (time savings, error rates, reuse metrics) and publish results to build the evidence base.
4. **Use AI/ML as assistive tools with full human oversight** (Evidence: Very Low; Recommendation: Conditional): LLM-generated code requires complete validation; treat as a “first draft” pending regulatory guidance. The gap between benchmark performance (88–93% on NLP tasks) and code generation accuracy (60–85%) suggests that domain-specific fine-tuning is necessary before clinical programming deployment.
5. **Build organizational competencies** in R/Python, version control, CI/CD, and metadata engineering— the convergence of data management and statistical programming requires expanded skill sets^2^.
6. **Document automation outcomes quantitatively**: organizations implementing new approaches should prospectively measure and publish efficiency metrics, error rates, and costs to build the evidence base that this review has shown to be deficient.

### 4.5 Limitations [PRISMA-ScR Item 19]

#### 4.5.1 Limitations of the Evidence Base

Critical limitations of the reviewed evidence include: (1) no RCTs exist comparing validation approaches (severity 100/100); (2) only 2.3% of validation papers provide quantitative data; (3) no longitudinal studies (≥2 years) track organizations transitioning to automated validation; (4) heterogeneous outcome definitions preclude cross-study comparison; (5) publication bias likely inflates reported efficiency gains; and (6) most evidence derives from large pharmaceutical companies, limiting generalizability to smaller organizations and academic medical centers.

#### 4.5.2 Limitations of This Review

This scoping review has several methodological limitations: (1) the protocol was registered retrospectively, which may introduce reporting bias; (2) inter-rater reliability for screening was not formally calculated; (3) the inclusion of conference proceedings and grey literature, while necessary given the field’s publication patterns, introduces heterogeneity in evidence quality; (4) the GRADE framework, designed for clinical interventions, was adapted for technical literature—this adaptation has not been externally validated; (5) the search was limited to English-language publications; and (6) as a scoping review, we mapped the evidence landscape rather than providing definitive estimates of effect sizes.

## 5 Conclusions [PRISMA-ScR Item 20]

This scoping review synthesized 262 publications (from 987 screened) on clinical trial statistical programming automation (2020–2025), applying GRADE evidence quality assessment to address three research questions. Key conclusions:

1. **RQ1: Technology is mature; evidence is not**. The pharmaverse ecosystem, metadata-driven architectures, and CDISC dataset automation tools have reached production maturity. However, only 16% of publications report quantitative outcomes, and most claims rest on Low to Very Low quality evidence. CDISC dataset automation (GRADE: Low) is the strongest evidence domain; TLF generation and validation claims (GRADE: Low–Very Low) should be interpreted cautiously. The Automation Maturity Framework (Figure 9) reveals an asymmetry between technology readiness and evidence quality.
2. **RQ2: Validation evidence is deficient**. Only 2.3% of 527 validation papers provide quantitative data, and just four studies yield directly comparable effectiveness data for specific approaches; no RCTs comparing validation approaches exist. Organizations spend 1.6–2.0×primary programming effort on double programming based on Very Low quality evidence. This is the most consequential gap identified, as it affects the most resource-intensive activity in the field.
3. **RQ3: A structured research agenda is needed**. We propose five prioritized studies (Table 5), led by a multi-site RCT comparing validation approaches (*n* ≥200 programs/arm) and a Delphi consensus process for standardized error detection metrics.

The Automation Maturity Framework provides organizations with a structured tool for assessing readiness across four dimensions and identifying targeted investments. By transparently reporting evidence quality alongside claimed benefits, this review enables evidence-informed rather than anecdote-driven adoption decisions. We urge organizations adopting automation to prospectively measure and publish outcomes, so that future reviews can provide stronger evidence-based guidance.

## Data Availability

All data produced in the present work are contained in the manuscript

## Acknowledgments

The authors acknowledge the contributions of the pharmaverse community, PHUSE, CDISC, and the R Validation Hub in advancing clinical programming automation standards and tools.

## Ethics Statement

This study was a literature review synthesizing published research and did not involve human subjects, patient data, or primary data collection. Therefore, ethics committee approval was not required for this study.

## Author Contributions

JY conceived and designed the review, conducted the literature search and screening, performed data extraction and synthesis, created all figures and tables, and wrote the manuscript. JZ contributed to title/abstract screening and verification of quantitative data extraction. TT verified a random 20% sample during full-text screening and contributed to manuscript revision. All authors approved the final version.

## Supplementary Materials

The following supplementary materials are available:

- **Table S1**: PRISMA-ScR 22-item checklist with page/line references
- **Table S2**: Full electronic search strategy for PubMed with MeSH terms and result counts
- **Table S3**: Complete data charting form template

