## Supplementart materials for "Evidence Behind the Automation of Clinical Trial Statistical Programming: A Scoping Review of Technology Adoption, Validation Frameworks, and AI/ML Integration (2020–2025)"

### Table S1: PRISMA-ScR (Preferred Reporting Items for Systematic Reviews and Meta-Analyses extension for Scoping Reviews) 22-Item Checklist

*Based on Tricco et al. (2018). Ann Intern Med; 169(7):467–473.*

| # | Section | Item | Reported | Location |
|---|---|---|---|---|
| *Title* | | | | |
| 1 | Title | Identify the report as a scoping review | Yes | Title, line 48 |
| *Abstract* | | | | |
| 2 | Abstract | Provide a structured summary that includes: background, objectives, methods, results, and conclusions | Yes | Lines 69–77 |
| *Introduction* | | | | |
| 3 | Rationale | Describe the rationale for the review in the context of what is already known. Explain why the review was conducted | Yes | Section 1.2 (lines 99–109) |
| 4 | Objectives | Provide an explicit statement of the questions being addressed with reference to participants, concept, and context (PCC) | Yes | Section 1.3 (lines 111–121) with PCC framework |
| *Methods* | | | | |
| 5 | Protocol and registration | Indicate whether a review protocol exists and if and where it can be accessed | Partial | Section 2.1; OSF DOI pending |
| 6 | Eligibility criteria | Specify characteristics of the sources of evidence used as eligibility criteria (PCC framework) | Yes | Section 2.2 (lines 130–139) |
| 7 | Information sources | Describe all information sources searched with dates of coverage | Yes | Section 2.3 (lines 141–145) |
| 8 | Search strategy | Present the full search strategy for at least one database, including any filters and limits used | Yes | Section 2.3; Supp Table S2 |
| 9 | Selection of sources of evidence | State the process for selecting sources of evidence (screening, eligibility, included) | Yes | Section 2.4 (lines 147–149) |

| | | | | |
|---|---|---|---|---|
| 10 | Data charting process | Describe the methods of charting data from included sources and any processes for obtaining or confirming data | Yes | Section 2.5 (lines 151–153) |
| 11 | Data items | List and define all variables for which data were sought | Yes | Section 2.6 (lines 155–167) |
| 12 | Critical appraisal of individual sources | Describe methods used for assessing critical appraisal of individual sources | Yes | Section 2.7 (lines 169–171); GRADE adaptation |
| 13 | Synthesis of results | Describe the methods of handling data and combining sources of evidence | Yes | Section 2.8 (lines 173–184) |
| *Results* | | | | |
| 14 | Selection of sources of evidence | Give numbers of sources screened, assessed for eligibility, and included, with reasons for exclusions | Yes | Section 2.4; Figure 1 (PRISMA flow diagram) |
| 15 | Characteristics of sources of evidence | For each source of evidence, present characteristics for which data were charted and provide citations | Yes | Table 5 (tab:characteristics) |
| 16 | Critical appraisal within sources of evidence | Present data on critical appraisal relevant to the review questions | Yes | Tables 2–4; Figures 5–6 (GRADE ratings) |
| 17 | Results of individual sources of evidence | For each result, present the data with citation | Yes | Sections 3.1–3.5 organized by RQ |
| *Discussion* | | | | |
| 18 | Synthesis of results | Summarize main results and provide a general interpretation with respect to the review questions | Yes | Section 4.1 (lines 846–855) |
| 19 | Limitations | Provide a general interpretation of the results and their implications for future research | Yes | Section 4.5 (lines 969–978) |
| *Funding* | | | | |
| 20 | Conclusions | Provide a general interpretation of the results and their implications | Yes | Section 5 (lines 981–989) |
| 21 | Funding | Describe sources of funding and role of funders | Yes | Funding section |
| 22 | Conflicts of interest | Declare any conflicts | Yes | Competing Interests section |

Table 1: PRISMA-ScR 22-item checklist with compliance status and manuscript location references. 21 of 22 items are fully compliant; Item 5 (protocol registration) is pending OSF DOI assignment.

### Table S2: Full Electronic Search Strategy for PubMed

*Search conducted January–March 2025 (Phase 1, core review) and April–June 2025 (Phase 2, validation meta-analysis). Last searched December 15, 2025.*

#### Phase 1: Core Review (January–March 2025)

| Search Step | Query | Results |
|---|---|---|
| *Block 1: Clinical Trial Context* | | |
| 1 | MeSH: “Clinical Trials as Topic” | 523,847 |
| 2 | “clinical trial” OR “clinical study” (title/abstract) | 892,341 |
| 3 | 1 OR 2 | 1,012,456 |
| *Block 2: Statistical Programming* | | |
| 4 | “statistical programming” OR “SAS programming” (title/abstract) | 2,156 |
| 5 | “CDISC” OR “SDTM” OR “ADaM” (title/abstract) | 4,823 |
| 6 | “TLF” OR “tables listings figures” (title/abstract) | 1,247 |
| 7 | 4 OR 5 OR 6 | 7,489 |
| *Block 3: Automation and Technology* | | |
| 8 | “automation” OR “automated” OR “metadata-driven” (title/abstract) | 387,654 |
| 9 | “validation” OR “double programming” (title/abstract) | 124,567 |
| 10 | “R programming” OR “pharmaverse” (title/abstract) | 3,891 |
| 11 | “artificial intelligence” OR “machine learning” OR “large language model” (title/abstract) | 234,512 |
| 12 | 8 OR 9 OR 10 OR 11 | 612,891 |
| *Combined Search* | | |
| 13 | 3 AND 7 AND 12 | 847 |
| 14 | Limit to 2020–2025 | 412 |
| 15 | Limit to English language | 398 |
| *Additional Sources* | | |
| 16 | Google Scholar (hand-searched, 2020–2025) | 289 |
| 17 | arXiv (cs.AI, cs.CL, 2023–2025) | 156 |
| 18 | Conference proceedings (PharmaSUG, PHUSE, R/Pharma, DIA) | 247 |
| 19 | Hand-searched reference lists and regulatory websites | 143 |
| **Total unique records after deduplication** | | **987** |

Table 2: PubMed search strategy with MeSH terms, Boolean operators, and result counts. Additional sources were searched concurrently and combined with PubMed results before deduplication.

#### Phase 2: Validation Meta-Analysis (April–June 2025)

An expanded search targeted validation-specific literature to support RQ2 analysis. The same database sources were used with modified search terms:

| Search Step | Query | Results |
|---|---|---|
| **Search Step** | **Query** | **Results** |
| 1 | “validation” AND (“clinical trial” OR “regulatory submission”) | 2,341 |
| 2 | “double programming” OR “independent programming” OR “peer review” | 891 |
| 3 | “risk-based validation” OR “risk-proportionate” | 423 |
| 4 | “automated testing” OR “snapshot testing” OR “CI/CD” AND “clinical” | 567 |
| 5 | “computer system validation” OR “21 CFR Part 11” OR “GAMP” | 1,289 |
| 6 | 1 OR 2 OR 3 OR 4 OR 5 | 3,891 |
| 7 | Limit to 2020–2025, English language | 1,647 |
| 8 | After deduplication with Phase 1 results | 527 |

Table 3: Phase 2 validation-focused search strategy. The 527 unique validation papers were screened for quantitative effectiveness data as reported in Section 3.2.

### Table S3: Data Charting Form Template

*The following form was developed in Microsoft Excel, pilot-tested on 15 publications, and iteratively refined before full data extraction.*

| Category | Data Item | Description / Instructions |
|---|---|---|
| *Bibliographic Information* | | |
| | Author(s) | First author last name, initials; et al. if > 6 authors |
| | Year | Publication year (4 digits) |
| | Title | Full article title |
| | Publication type | Journal / Conference / Guidance / Documentation |
| | Journal/Proceedings | Full name of journal or conference |
| | Country/Region | Country of first author’s affiliation |
| *Methodological Characteristics* | | |
| | Study design | RCT / Controlled evaluation / Case study / Before-after / Expert opinion / Survey |

| | | |
|---|---|---|
| | Sample size | Number of programs, datasets, or participants evaluated |
| | Comparator | Control group or baseline method (if applicable) |
| | Study duration | Time period covered (if longitudinal) |
| | Multi-site? | Yes/No; number of sites if Yes |
| *Domain and Technology* | | |
| | Primary domain | TLF generation / CDISC automation / Validation / AI/ML / Platform |
| | Secondary domain(s) | Additional domains addressed (if any) |
| | Tools/platforms | Specific tools or packages described (e.g., rtables, admiral, GPT-4) |
| | Programming language(s) | SAS / R / Python / Other |
| | CDISC standard version(s) | SDTM IG version, ADaM IG version, ARS version (if applicable) |
| *Outcomes and Metrics* | | |
| | Quantitative outcomes reported? | Yes/No |
| | Time reduction (%) | Reported range or point estimate with measure of variance |
| | Error detection rate (%) | Reported rate with denominator |
| | F1/Accuracy score | For AI/ML evaluations |
| | Cost savings | Reported monetary savings or FTE reductions |
| | Variance estimates | SD, SE, CI, or $p$-value if reported |
| | Qualitative findings | Key themes, lessons learned, barriers identified |
| *Evidence Quality Assessment* | | |
| | GRADE rating | High / Moderate / Low / Very Low |
| | GRADE justification | Specific reason for rating (study design, sample, comparator) |
| | Publication bias concern | Low / Moderate / High (with rationale) |
| | Generalizability assessment | Limited / Moderate / Broad (with rationale) |
| *Extraction Metadata* | | |
| | Extracted by | Reviewer initials |
| | Extraction date | DD/MM/YYYY |
| | Verification status | Pending / Verified by second reviewer / Discrepancy resolved |

| | |
|---|---|
| Notes | Any additional observations or queries |

Table 4: Complete data charting form template used for extraction from the 262 included sources. The form was iteratively refined during pilot testing on 15 publications.
